# Clinical Performance and Safety of Cerviron Vaginal Ovules in the Management of Cervical Lesions Postoperative Care: A National, Multicentric Study

**DOI:** 10.1101/2023.10.16.23297062

**Authors:** Izabella Petre, Daniela Oana Toader, Ramona Petrita, Alexandru-Remus Pinta, Andreea Anda Alexa, Marina Adriana Mercioni, Roxana-Mihaela Sandulescu

## Abstract

**Background:** Laser conization and electrocautery are the preferred surgical options for the management of benign cervical erosions. Local re-epithelialization therapy should be initiated preoperatively and postoperatively to prevent severe bleeding, wound inflammation, and infection.

**Objective:** To investigate the aspect of the cervix by colposcopy after a 3-month treatment with the medical device. As secondary objectives, the study analyzed the presence of primary and secondary inflammation, the presence of ulceration, colpitis, vaginal discharge, and vaginal irritation.

**Methods:** The study population included 27 participants with benign, ectopic, cervical ectopy that consented their participation in 3 research sites from Romania. The treatment protocol consisted of the delivery of the medical device intravaginally, for 15 consecutive days, with a total study duration of 3 months.

**Results:** Cerviron had a positive impact on cervical epithelialization. After 3 months of treatment with Cerviron, for 100.00% of the participants, the colposcopy result was normal. Cerviron seems to exert a beneficial effect in reducing primary inflammation correlated with cervical erosions. It was observed that after 90 days of treatment primary inflammation was reduced by 85.19% (p < 0.001; 95% CI [58.74 – 98.35]). Other clinical parameters associated with cervical ectopy have certainly improved: vaginal erosion by 70.37% (p < 0.001, 95% CI [48.30 – 93.23]), ulceration by 55.56% (p < 0.001, 95% CI [33.22 – 83.67]), colpitis by 81.48% (95% CI [62.52 – 100.00], and vaginal secretion by 66.67% (95% CI [44.26 – 90.74]).

**Conclusions:** The medical device reduced inflammation and other symptoms related to non-malignant cervical disease. Moreover, our study findings suggest that supportive treatment with Cerviron can be recommended for cervical wound healing in patients with human papillomavirus infection.

ClinicalTrials.gov identifier: NCT04735718

## Introduction

### Background

The cervix is a muscular organ covered by two types of mucous membranes (epithelia), namely: a squamous epithelium (consisting of several overlapping layers which, as they become mature, detach and are found in the secretion around the cervix) and a cylindrical epithelium or glandular (consisting of a single layer of mucus-secreting cells). The two types of epithelia are continuous with each other and thus cover the surface of the endocervical canal. The place where the two types of epithelia meet is called the “squamous-cylindrical junction”. The proportion between the two epithelia mentioned above varies physiologically between puberty and post-menopause so the position of the junction also varies with age. The cervix acts like a barrier to prevent bacteria and viruses from entering the uterus [1]. When the cervix is infected, there is an increased risk for the infection to spread into the uterus [2]. The squamous-cylindrical junction is the main site through which the Human papillomavirus (HPV) enters the epithelium and can infect its deep cells. The HPV infection is correlated with cervical erosions, as well as other pathologic conditions such as benign neoplasms, premalignant, malignant, or congenital anatomic anomalies) [3][4]. The treatment of cervical erosions has an important significance for preventing the occurrence of precancerous lesions and cervical cancer [5]. A plethora of symptoms such as vaginal discharge, vulvar itching, dyspareunia, pelvic pain, postcoital bleeding, nocturia, and frequency of micturition are associated with cervical erosion [6,7].

### Benign Cervical Ectopy – the decision to treat

The cervix can present a variety of pathologies that include benign entities (such as cervical ectropion, cervical polyps, genital warts, myomas, Nabothian cysts, or cervical hypertrophy) as well as malignant lesions, the most common being cervical carcinomas [8]. Normally, the cervix is covered with squamous epithelial cells on its outer surface and contains glandular cells within the cervical canal. However, in cervical ectropion, these glandular cells are found on the outer surface of the cervix, where they are not typically present.

Prevalence studies show that cervical ectopy (CE) can vary between 14-54.9% and it increases with parity [8][9][10]. A multitude of cervical lesions appear as a result of exposure to estrogen (such as contraceptive pills) or more controversial, infection with HPV [11][12][13][14]. Erosion was significantly more common in women taking oral contraceptives and less common in women using barrier methods of contraception [9].

Identifying and treating CE, as well as options for appropriate treatment, are current challenges for medical specialists. While several studies reveal different therapeutic approaches, the decision to treat or not remains controversial [15–17]. To our knowledge, few studies, if any, are focusing on the postoperative management of cervical wounds in the Western population.

The objective of our study was to verify the performance of the medical device Cerviron when applied after surgical intervention for the treatment of benign CE. The device’s effects in non-specific vaginitis were confirmed in a recent study [18]. For 72.34% patients treated with Cerviron^®,^ the medical device had a beneficial effect on the score of vaginal symptoms. It was observed that the treatment for 90 days with Cerviron^©^ vaginal ovules significantly reduced the following symptoms: leucorrhea (*p < 0*.*001*), pruritus (*p < 0*.*05*), burn (*p < 0*.*05*), rash (*p < 0*.*05*), pain *(p < 0*.*001*), malodor (*p < 0*.*05*), dysuria (*p < 0*.*05*), and dyspareunia (*p < 0*.*001*).

## Materials and methods

### Study Design

This study was designed as an open-label, pilot, multicentric, national clinical investigation. The study took place between July 09, 2021 (first participant in) and January 6, 2023 (last participant out), in 3 investigational sites located in Romania. The selected sites were two county hospitals (Institutul National pentru Sanatatea Mamei si Copilului “Alessandrescu-Rusescu” - Department of Obstetrics-Gynecology III, Principal Investigator Dr. Daniela Oana Toader and Emergency County Clinical Hospital “Pius Brinzeu” Timisoara - Department of Obstetrics-Gynecology II, Principal Investigator Dr. Izabella Petre) and one private practice (Misca Medical Center Timisoara, Principal Investigator Dr. Flavius Olaru).

The purpose of this clinical investigation was to confirm the therapeutic performance and tolerability of an intravaginal medical device in postoperative cervical lesions. The effects were measured as differences in clinical performance determined by colposcopy results and the existence of vaginal symptoms assessed by the participant between the baseline visit and three successive study visits (scheduled at 30-day intervals). The evaluation of the medical device’s performance comprised the following clinical outcomes: change in vaginal discharge aspect, change in vaginal pH values, change in vaginal microbiome, change in vaginal inflammation, performed every 30 days, as shown in figure 1.

**Figure 1.**
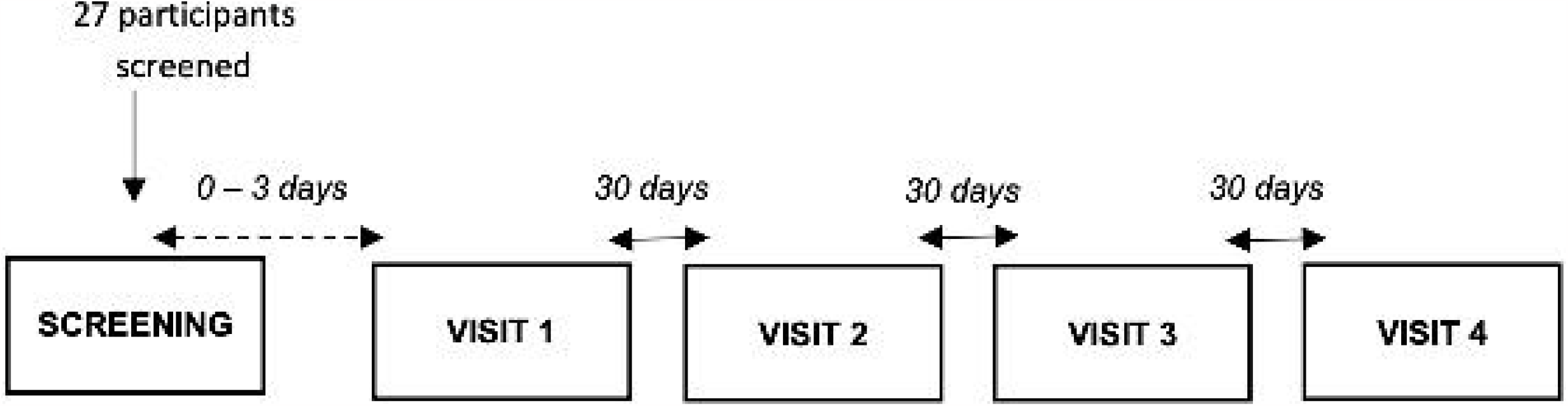
Study Flow Chart

### The Medical Device

Cerviron is a medical device indicated as an adjuvant treatment in atrophic, irritant, and inflammatory vaginitis caused by the imbalance of the vaginal pH and changes in the vaginal microflora. The device is formulated as vaginal ovules with a complex composition, as follows: bismuth subgallate 100 mg, hexylresorcinol 2 mg, vegetable collagen 15 mg, *Calendula officinalis* 10 mg, *Hydrastis canadensis* 10 mg, *Thymus vulgaris* 10 *mg, and Curcuma longa* 10 mg). The instructions for use specify its field of use as an adjuvant in the treatment of acute and chronic vulvovaginitis and cervical lesions of mechanical origin. The recommended use of the device is 1 ovule per day inserted on the first day after the menstruation. Treatment should be continued for 15 consecutive days. Its adjuvant role in the treatment of non-specific vaginitis was outlined in a recent clinical investigation (NCT04735705) and a real-world study (NCT05668806) revealed its intended use as a treatment for cervical lesions of different etiologies [18][19]. It favors the healing and re-epithelialization processes and reduces the proliferation of endogenous pathogens. The medical device is indicated for the treatment of vaginal symptoms such as leukorrhea, vaginal pruritus, burns in the vaginal area, rash, vaginal discharge with malodor, dysuria, dyspareunia, and vaginal bleeding. The study architecture is presented in figure 2.

**Figure 2.**
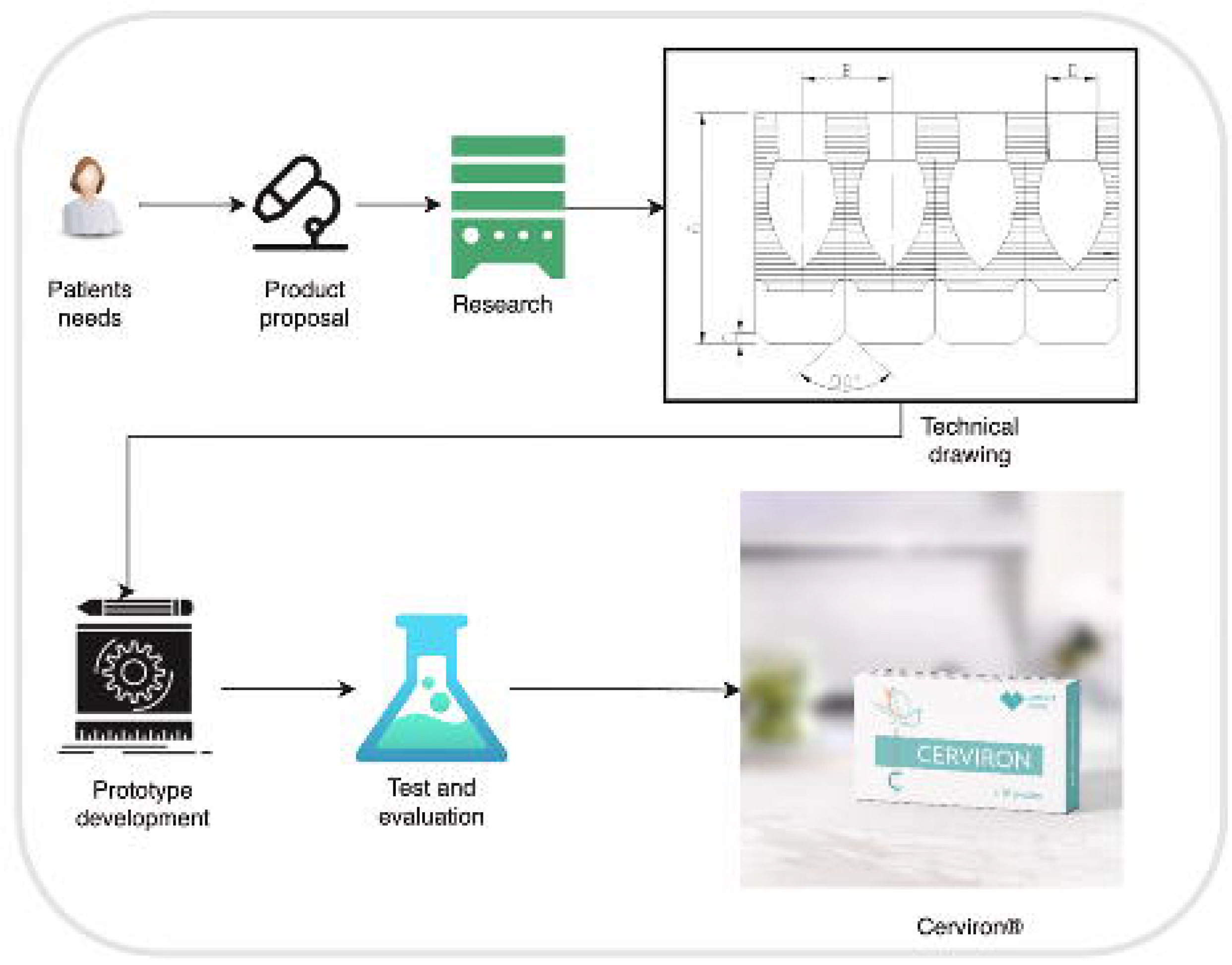
The study architecture

### Participant Population

The target population for this clinical investigation was comprised of women aged 18 to 65 years presenting a normal cervical cytology report, e.g., Negative for Intraepithelial Lesions or Malignancy (NILM), (LSIL), atypical glandular cells or slightly modified to Atypical Squamous Cells of Undetermined Significance (ASC-US) in the last 6 months, that underwent surgical removal of benign, cervical lesions. Vulnerable subjects were not enrolled in this clinical investigation. Vaginal tampon use was prohibited during the clinical investigation because tampons can prevent the ovule from exhibiting its full performance. In addition, a list of restricted concomitant medications was defined in the clinical investigation plan to distinguish between the efficacy of the medical device in question and the efficacy of other therapies. All concomitant medications and treatments were recorded in the appropriate study documents. Colposcopy examination with LEO-2100I was performed at baseline and following the 3 consecutive months the medical device was applied. A colposcopy view of the cervix was performed using Lugol’s iodine.

Adverse events and concomitant medications were collected at each visit. Each participant was followed up for 3 months or approximately 90 days. The data were exported and analyzed using Analyse-It for Mi-Microsoft Excel, version 5.90 (Analyse-It Software Ltd, Leeds, UK).

### Data Collection

The clinical investigation included subjects under treatment with Cerviron vaginal ovules for approximately 3 months (Fig. 1). Each enrolled subject performed 4 visits. The screening visit (SV) took place 3-0 days before the baseline visit (day 0). On day 0 the subjects received the medical device. The study participants had 4 visits to the investigational site, at 30-day intervals each.

A total number of 27 participants were screened during the enrolment period of 15 months, between 12 July 2021 and 18 October 2022. Out of 27 subjects enrolled in the clinical study, one subject (0207) did not meet the inclusion and exclusion criteria, as a result, she was not included in the clinical trial and one subject (0307) was lost to follow-up. The safety population included the 25 subjects who took the treatment. All subjects had a 100% compliance. The Intention-to-Treat population included 26 subjects who received the treatment of the study medical device. The Per Protocol Set population, subjects who completed all the visits without major protocol deviations, was 25 subjects.

The National Ethics Committee (CNBMDM) evaluated all participant-related documentation and released its favourable opinion before the site initiation. Initial Approval 2DM was received on 01 April 2021 (protocol number CYRON/02/2021; NCT04735718). The clinical data were gathered and assessed by the Helsinki Declaration’s ethical criteria. Before any research-specific procedure, participants’ informed consent forms for inclusion in the study were signed.

### Statistical Analysis

Statistical analysis and generation of tables, figures, and patient data listings were performed using R statistical software version 4.1.1 (R Foundation for Statistical Computing, Vienna, Austria) with a few extra statistical packages added, all of which have been re-visited and updated to the latest version. The final analysis was completed after all participants finalized the study, all queries were solved, and the database was locked. The overall type I error was preserved at 5%; P < 0.05 was considered statistically significant, and 95% CIs were calculated.

To evaluate changes of proportions over time before and after the treatment, for categorical variables, a two-proportions z-test or Fisher Exact Test was performed. The change in vaginal score between visits was evaluated using the Kruskall-Wallis test and for detecting the change in vaginal score for the first 30 days of treatment Wilcoxon signed-rank test was performed.

The following primary results were collected in this study: colposcopy examination which may be 1 - Normal or 2 - Abnormal, the presence of primary and secondary inflammation, the presence of erosions, ulcers, colpitis, vaginal discharge, and vaginal irritation.

Success was defined by resolution (normal colposcopy result) and substantial improvement in clinical signs after 90 days of treatment with Cerviron.

As secondary outcomes, the following clinical symptoms and signs were verified to demonstrate the efficacy and performance of Cerviron: re-epithelialization degree (by visual examination), primary inflammation, secondary inflammation, erosion, ulceration, colpitis, vaginal discharge, and vaginal irritations.

A typical satisfaction survey (5-point Likert Scale) was used as a measurement of participant satisfaction and comfort.

## Results

Of the 26 participants initially enrolled, 25 patients completed the study. All 25 patients had an abnormal colposcopy examination (100.00%), and for 0.00% (N = 0) participants the colposcopy examination was normal. During the colposcopy examination, at the screening visit, participants presented cervical erosions, ulcerations, and colpitis macularis (“*strawberry cervix*”). Three patients have had Nabothian cysts and four participants had an associated HPV infection. Primary inflammation was present for all study participants. Table 1 shows the demographic data of all study participants.

**Table 1.** The demography of the study participants.

Success was defined by resolution (normal colposcopy result) and substantial improvement in clinical signs after 90 days of treatment with Cerviron. Results are shown in table 2.

After 3 months of treatment with Cerviron, for 100.00% of the participants (N=25) the colposcopy was normal, while no patient presented an abnormal result (N=0). (p < 0.001; 95% CI [96.00 – 100].

At the same time, the following clinical symptoms were verified to demonstrate the efficacy and performance of Cerviron. Thus, at the first visit, 96.00% of the study participants (N = 24) presented primary inflammation, 84.00% (N = 21) erosion, 68.00% (N = 17) vaginal ulcerations, 92.00% (N = 23) colpitis, and 80.00% (N = 20) vaginal secretions. Table 3 shows all the clinical symptoms that were investigated to determine the effectiveness of the medical device.

It can be observed that after 90 days of treatment primary inflammation, vaginal erosion, ulceration, colpitis, and vaginal secretion were reduced by 100%.

The effect of the medical device on different vaginal symptoms was verified. Thus, at the first visit, 96.00% of the study participants (N = 24) presented primary inflammation, 84.00% (N = 21) erosion, 68.00% (N = 17) vaginal ulcerations, 92.00% (N = 23) colpitis, and 80.00% (N = 20) vaginal secretions (Table 4).

### Degree of re-epithelialization

The degree of epithelialization was assessed every 30 days (at each visit) using a score as follows: 1-Absent, 2 - Partial, and 3 - Complete. The change in the degree of epithelialization during visits 1 - 4 is shown in table 4.

**Table 4.** Degree of re-epithelialization between each visit.

At the baseline visit, the degree of epithelialization was absent for 18 patients (72.00%), while for 7 patients (28.00%) it was partial. No patients had a complete degree of epithelization. After 30 days of treatment with Cerviron, it was observed a significant improvement in the degree of epithelialization as follows: 24 patients (96%) had a partial degree of epithelization, and 1 patient (4.00%) had a complete degree (p < 0.001, Fisher’s Exact Test). Moreover, after 60 days of treatment, Cerviron’s performance is more pronounced. Thus, at 60 days of treatment, several 3 patients (12.00%) had a partial degree of epithelization, and 22 patients (88.00%) had a complete degree (p < 0.001, Fisher’s Exact Test). At the end of the treatment schedule, after 90 days, all patients presented a complete degree of epithelialization, which shows that Cerviron^®^’s performance was 100% as expected (p < 0.001, Fisher’s Exact Test).

### Change in vaginal symptoms

The secondary objective of the study was to evaluate the evolution of symptomatology during the treatment with Cerviron.

For evaluation of the efficacy of the medical device were assessed the vaginal symptoms score such as bleeding, malodor, dysuria, dyspareunia, pain, and leucorrhea on a scale of 0 to 5, where 0 represents no intensity and 5 represents the maximum intensity of symptoms, as shown in table 5.

**Table 5.** Means and Standard Deviations of each symptom between visits.

Thus, it was observed that treatment with Cerviron reduces bleeding score after 90 days in all cases from a mean score of 1.44 ±1.16 points at baseline to 0.00 ± 0.00 points at the final visit (p < 0.001), malodor score after 90 days in all cases from a mean score of 1.52 ± 1.33 points at baseline to 0.00 ± 0.00 points at final visit (p < 0.001), dysuria symptoms from a mean score of 1.40 ± 1.35 points at baseline to 0.20 ± 1.00 points at final visit (p < 0.001), dyspareunia symptoms from a mean score of 1.68 ± 1.57 points at baseline to 0.00 ± 0.00 points at final visit (p < 0.001), pain score after 90 days in all cases from a mean score of 1.60 ± 1.32 points at baseline to 0.00 ± 0.00 points at final visit (p < 0.001), and leucorrhea symptoms after 90 days in all cases from a mean score of 2.08 ± 1.38 points at baseline to 0.00 ± 0.00 points at final visit (p < 0.001).

Treatment with Cerviron shows significant efficacy (Figure 3a) in reducing bleeding score after 30 days with 61.11% from 1.44 ± 1.16 at baseline to 0.56 ± 0.71 (p < 0.05). Also, it shows significant efficacy to reduce malodor score with 63.16% (Figure 3b) from 1.52 ± 1.33 points to 0.56 ± 0.71 points (p < 0.05), dysuria with 42.86% (Figure 3c) from 1.40 ± 1.35 point to 0.80 ± 1.19 points (p = 0.083), dyspareunia score with 59.52% (Figure 3d) from 1.68 ± 1.57 to 0.68 ± 0.85 (p < 0.05), pain score with 65.00% (Figure 3e) from 1.60 ± 1.32 points to 0.56 ± 0.65 points (p < 0.05), and leucorrhea symptoms with 73.08% (Figure 3f) from 2.08 ± 1.38 points to 0.56 ± 0.65 points (p < 0.05).

**Figure 3.**
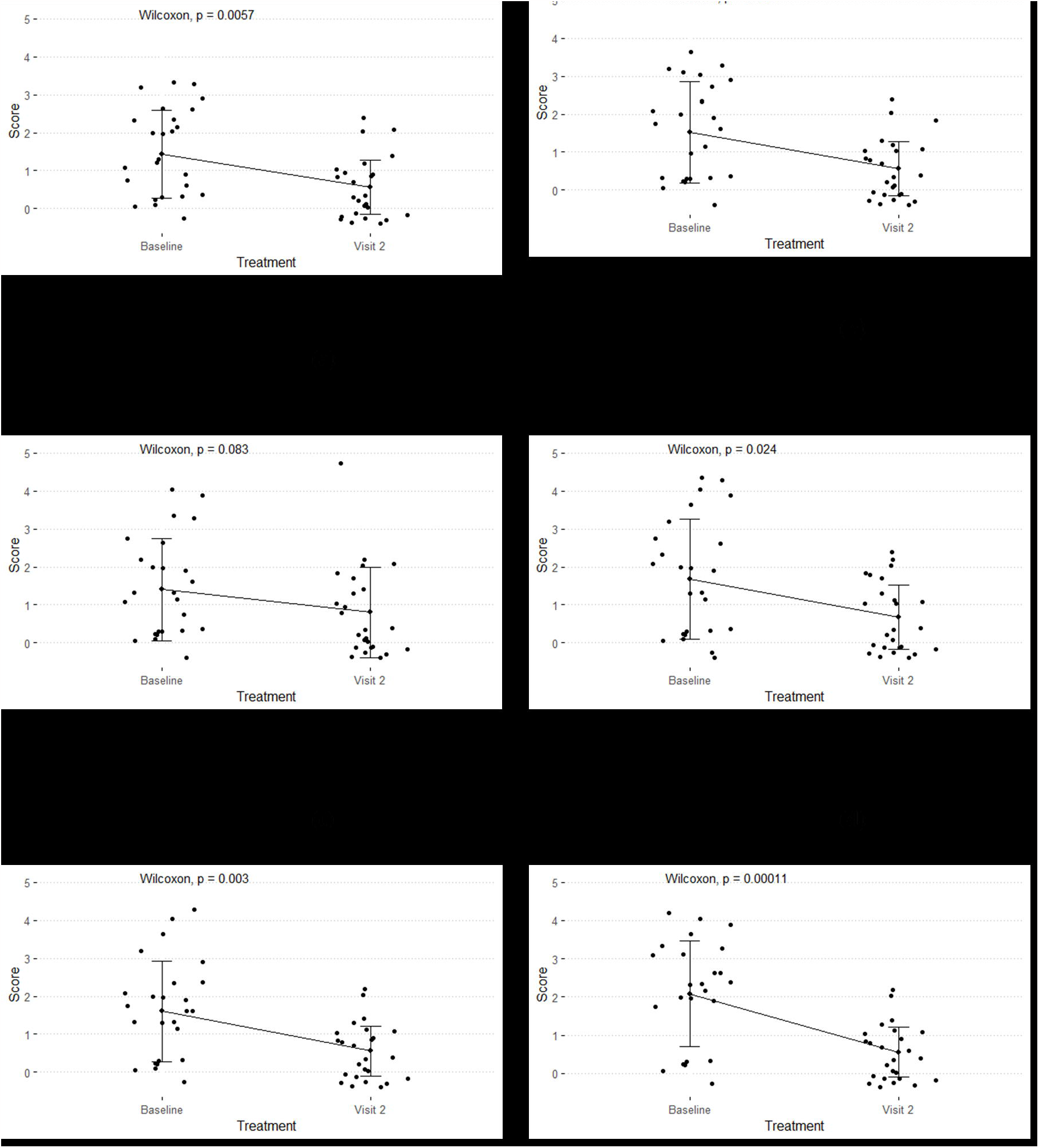
Improvement score after 30 days of treatment with Cerviron (**a**) in bleeding; (**b**) in malodor; (**c**) in dysuria; (**d**) in dyspareunia; (**e**) in pain; (**f**) in leucorrhea.

**Figure 4.**
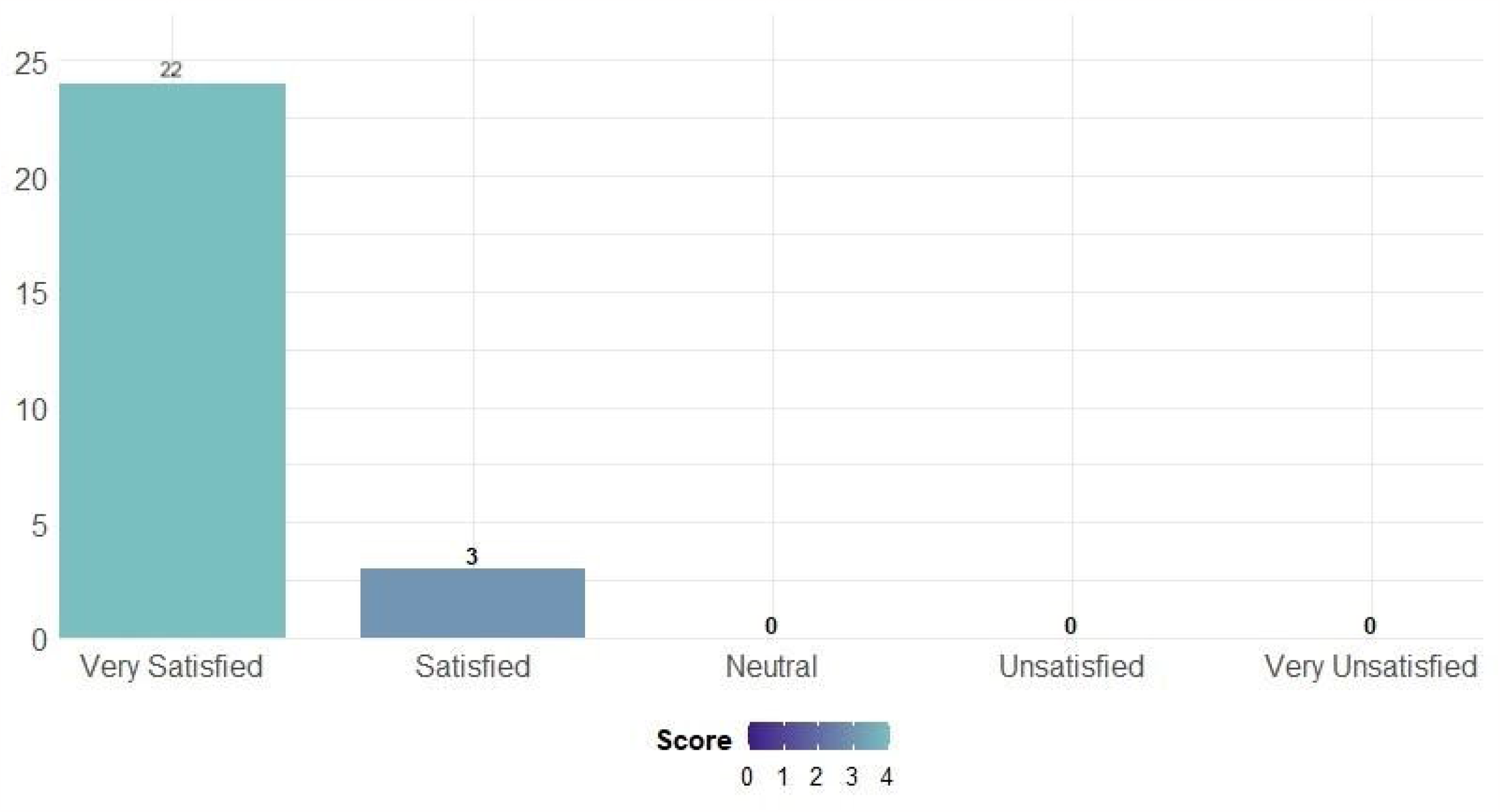
Participant Satisfaction Score – Likert Scale.

The degree of satisfaction when using the medical device was assessed using a five-point Likert Scale, as very satisfied, satisfied, neutral, unsatisfied, and very unsatisfied. The evaluation of participant satisfaction was summarized in the following figure. The majority of participants (88.00%) in the study declared themselves very satisfied (N=22), while 12.00% (N=3) were satisfied with the product.

## Discussion

### Reducing primary inflammation associated with cervical ectopy

Some factors involved in the pathogenesis of CE involve the action of estrogens [20]. Estrogens influence the inflammatory processes, by regulating chemokines and chemokine receptors. Straub and colleagues studied the complex processes of inflammation related to estrogen signaling toward immune cell trafficking [21]. The T-helper 17 cells producing interleukins are the main T cells responsible for chronic inflammation. As such, conditions with high estrogen exposure, such as adolescents, pregnancy, the use of hormonal contraception are commonly linked to the appearance of cervical ectropion [12]. There are fewer CD3+, CD4+, and CD8+ cells in cervical lesions compared to normal epithelium [22]. A correlation was found between cervical ectropion and the development of desquamative inflammatory vaginitis with overlapping symptoms [23].

An important finding of this study is that the medical device seems to exert a beneficial effect in reducing primary inflammation correlated with CE. Also, other clinical parameters associated with CE such as vaginal erosion, ulceration and leukorrhea production have certainly improved. As non-malignant cervical disease is correlated with female sexual dysfunction, these findings support and extend its role in improving sexual function in these patients, by reducing dyspareunia and dysuria [24]. Further research is needed to ascertain its use in these patients.

### The risks of the HPV infection and cellular reprogramming

Metaplasia tends to occur in tissues constantly exposed to environmental agents, which are often injurious in nature. Cervix metaplasia occurs when exposed to certain stimuli, such as low vaginal pH and HPV. An association between ectopy and HPV (OR = 2.09; p = 0.005) was found in a cross-sectional study, conducted by Rocha-Zavaleta. According to the study results, the researchers found a higher general HPV rate and a higher rate of HPV-16 in women with ectopy [25]. The study by Sarkar and Steele suggested that there was an association between cervical intraepithelial neoplasia (CIN) and ectopy [26]. Universally, metaplasia is a precursor to low-grade dysplasia, which can culminate in high-grade dysplasia and carcinoma [27]. Moreover, histopathological diagnosis shows that postcoital bleeding is often associated with CIN, cervical polyps, and inflammatory cervical smears bearing a high risk for invasive cervical cancer [28].

Still, in debate among specialists, routine treatment in CE is recommended, especially when considering taking preventive action against cervical cancer. Ectopy is modified over time by squamous metaplasia and epithelialization, low pH, trauma, and possibly by cervical infection. There is a relationship between squamous metaplasia and induction of squamous cell carcinoma of the cervix. Cells undergoing metaplasia are more susceptible to carcinogens. Precancerous lesions often develop at the squamous-columnar junction, i.e., the area of transition between glandular and stratified epithelium, which is the location where metaplasia is most intense. Also, cervical cysts left untreated can induce considerable enlargement of the cervix, which can lead to symptomatology. Complications of Nabothian cysts include hematometra, labor passage obstruction, rectal compression, abnormal uterine bleeding, specifically in the case of giant cysts, and chronic urinary retention by restricting the bladder’s outlet or by compressing the pudendal and sacral nerves, thus, disturbing the nerve supply to the detrusor muscle. Cerviron seems to have a very potent action in rebalancing the vaginal pH. In a previous clinical investigation, NCT04735705, vaginal pH values measured over 90 days showed that Cerviron restores altered vaginal pH [18].

Through its ingredient, bismuth subgallate, Cerviron ovules exert a local protective action on mucous membranes and raw surfaces [29,30]. Moreover, it contains collagen with nutritive, hydrating, healing, and trophic effects. Collagen is a key protein favoring the forming the scars during the healing of conjunctive tissues, due to its chemotactic role. Many collagen bandages were developed to improve the repair of the wound, especially of non-infected, chronic, idle cutaneous ulcerations. The Marigold extract (Calendula Officinalis) exerts the role of preventing fatty base degradation.

### Prevention of infections with sexually transmitted microorganisms such as Chlamydia Trachomatis

Recognition of CE should alert the clinician to the possibility of a genital *Chlamydia* infection. Opportunistic screening for Chlamydia in young people should be offered to reduce the prevalence of infection and its sequelae. *Chlamydia trachomatis* is the most common bacterial sexually transmitted disease associated with cervical ectropion [6,31]. In women, chlamydial infection usually presents as cervicitis, which can lead to up to 30% to 50% of pelvic inflammatory disease episodes. Pelvic inflammatory disease has significant reproductive sequelae, such as tubal infertility and ectopic pregnancy [32]. Out of 26 participating subjects, there were 0 cases of infection involving sexually transmitted microorganisms such as *Chlamydia trachomatis*. As such, Cerviron supportive therapy may be prescribed in sexually active patients for at least 1-3 months, 10-15 consecutive days, to prevent these infections.

Estrogens are capable of dramatically altering the responses of host cells to microbes. In adolescence, pregnancy, during hormonal contraception, or the years of menstruation (mostly in the ovulatory phase), the probability of developing CE is very high, and sometimes it goes undetected. Furthermore, CE implies further risks of acquiring STDs (Gonorrhea, Chlamydia, and HPV). When adding Cerviron either as monotherapy or in association with other antimicrobials in postoperative care, we observed better postoperative outcomes. The study results show that a 45-days treatment (3 treatment sessions of 15 days each) with Cerviron vaginal ovules was beneficial in providing a complete degree of cervical epithelialization and reduced the multitude of vaginal symptoms, including bleeding, inflammation, malodor, dysuria, dyspareunia, pain, and leucorrhea. This result was consistent with previous clinical studies that included the same medical device, NCT04735705, and NCT05668806 [18,19].

### Efficacy in patients with Intrauterine Contraceptive Devices (IUCD)

IUCD usage could be linked to inflammation, trauma of the cervix and increased vaginal discharge [33]. Cerviron should be recommended in patients using IUCD to reduce vaginal discharge, reduce primary inflammation, and to improve the epithelialization of the cervical area.

The particular limitations of the study include the absence of a control group, the small sample size, and a limited timeframe of only 3 months. Larger studies to include a more diverse population should be designed to validate the conclusions of our study.

### Study limitations

The particular limitations of the study include the absence of a control group, the small sample size, and a limited timeframe of only 3 months. Larger studies to include a more diverse population should be designed to validate the conclusions of our study.

## Conclusion

Cerviron ovules are an excellent adjuvant for the acceleration of cervical erosion healing. It favors the re-epithelialization of the damaged tissue and restoration of the initial colpo-ecosystem. Its topical application was observed to be effective in minimizing inflammation after surgical procedures and also in reducing unpleasant symptoms such as vaginal discharge, vaginal bleeding, malodor, dysuria, and dyspareunia.

Cerviron shows curative effects that support its use in enhancing cervical epithelialization after laser conization and electrocautery procedures. These findings suggest its use in the management of non-malignant cervical disease, including cervicitis, CIN, cervical polyps, Nabothian cysts, cervical hypertrophy, or cervical genital warts. Moreover, our study findings suggest that supportive treatment with Cerviron can be recommended for cervical wound healing in patients with HPV infection. In terms of safety, the medical device proved safe, as no study participant reported treatment-related AEs or SAEs. The only contraindication to Cerviron ovules has been determined as hypersensitivity to the active substance or any of the excipients within the medical device.

## Data Availability statement

Data will be made available on request.

## Acknowledgments

The authors thank the study participants who voluntarily enrolled in the study. The authors also thank Florentina Calancea for the technical support provided during data management collection and to Perfect Care Distribution for providing a research grant.

Author Contributions

All authors have equal contribution in manuscript review, editing, data curation, methodology, project administration and the interpretation of the study results. All authors approved the final version of the manuscript to be published.

## References

[1] De Tomasi JB, Opata MM, Mowa CN. Immunity in the Cervix: Interphase between Immune and Cervical Epithelial Cells. J Immunol Res 2019;2019:1–13. 10.1155/2019/7693183.

[2] Jain MA, Limaiem F. Cervical Squamous Cell Carcinoma. StatPearls, Treasure Island (FL): StatPearls Publishing; 2023.

[3] Lu Z, Chen H, Zheng X-M, Chen M-L. Expression and clinical significance of high risk human papillomavirus and invasive gene in cervical carcinoma. Asian Pac J Trop Med 2017;10:195–200. 10.1016/j.apjtm.2017.01.007.

[4] Casey PM, Long ME, Marnach ML. Abnormal cervical appearance: what to do, when to worry? Mayo Clin Proc 2011;86:147–50; quiz 151. 10.4065/mcp.2010.0512.

[5] Merera D, Jima GH. Precancerous Cervical Lesions and Associated Factors Among Women Attending Cervical Screening at Adama Hospital Medical College, Central Ethiopia. Cancer Manag Res 2021;13:2181–9. 10.2147/CMAR.S288398.

[6] Matiluko AF. Cervical ectropion. Part 2: assessment of symptomatic women. Trends Urol Gynaecol Sex Health 2009;14:16–22. 10.1002/tre.115.

[7] Agah J, Sharifzadeh M, Hosseinzadeh A. Cryotherapy as a Method for Relieving Symptoms of Cervical Ectopy: A Randomized Clinical Trial. Oman Med J 2019;34:322–6. 10.5001/omj.2019.63.

[8] Aggarwal P, Ben Amor A. Cervical Ectropion. StatPearls, Treasure Island (FL): StatPearls Publishing; 2023.

[9] Goldacre MJ, Loudon N, Watt B, Grant G, Loudon JD, McPherson K, et al. Epidemiology and clinical significance of cervical erosion in women attending a family planning clinic. BMJ 1978;1:748–50. 10.1136/bmj.1.6115.748.

[10] Cotarcea S, Stefanescu C. The Importance of Ultrasound Monitoring of the Normal and Lesional Cervical Ectropion Treatment. Curr Health Sci J 2016:188–96. 10.12865/CHSJ.42.02.11.

[11] Hwang LY, Lieberman JA, Ma Y, Farhat S, Moscicki A-B. Cervical Ectopy and the Acquisition of Human Papillomavirus in Adolescents and Young Women: Obstet Gynecol 2012;119:1164–70. 10.1097/AOG.0b013e3182571f47.

[12] Aggarwal P, Ben Amor A. Cervical Ectropion. StatPearls, Treasure Island (FL): StatPearls Publishing; 2023.

[13] for the Ludwig-McGill Cohort Study, Shaw E, Ramanakumar AV, El-Zein M, Silva FR, Galan L, et al. Reproductive and genital health and risk of cervical human papillomavirus infection: results from the Ludwig-McGill cohort study. BMC Infect Dis 2016;16:116. 10.1186/s12879-016-1446-x.

[14] Cao X, Gao N, Huang L, Yao J. Correlation of subclinical HPV infection with genital warts and cervical erosion. Eur J Gynaecol Oncol 2013;34:462–5.

[15] Pegu B, Srinivas BH, Saranya TS, Murugesan R, Priyadarshini Thippeswamy S, Gaur BPS. Cervical polyp: evaluating the need of routine surgical intervention and its correlation with cervical smear cytology and endometrial pathology: a retrospective study. Obstet Gynecol Sci 2020;63:735–42. 10.5468/ogs.20177.

[16] Shallal F. Clinical observation of electrocauterization alone and post-electrocauterization MEBO application therapy in the treatment of cervical erosion. Mustansiriya Med J 2015;14:7.

[17] Sakhipova GZ, Abenova NA, Zhumabaeva TN, Seypenova AN, Shamshaeva DO. Clinical Experience in Management of Patients with Cervical Erosion and Ectopia. Open Access Maced J Med Sci 2020;8(B):226–30.

[18] Toader DO, Olaru RA, Iliescu D-G, Petrita R, Calancea FL, Petre I. Clinical Performance and Safety of Vaginal Ovules in the Local Treatment of Nonspecific Vaginitis: A National, Multicentric Clinical Investigation. Clin Ther 2023;0. 10.1016/j.clinthera.2023.06.023.

[19] Petre I, Sirbu DT, Petrita R, Toma A-D, Peta E, Dimcevici-Poesina F. Real-world study of Cerviron^®^ vaginal ovules in the treatment of cervical lesions of various etiologies. Biomed Rep 2023;19:1–9. 10.3892/br.2023.1618.

[20] Machado Junior LC, Dalmaso ASW, Carvalho HBD. Evidence for benefits from treating cervical ectopy: literature review. Sao Paulo Med J 2008;126:132–9. 10.1590/S1516-31802008000200014.

[21] Straub RH. The Complex Role of Estrogens in Inflammation. Endocr Rev 2007;28:521–74. 10.1210/er.2007-0001.

[22] Litwin TR, Irvin SR, Chornock RL, Sahasrabuddhe VV, Stanley M, Wentzensen N. Infiltrating T-cell markers in cervical carcinogenesis: a systematic review and meta-analysis. Br J Cancer 2021;124:831–41. 10.1038/s41416-020-01184-x.

[23] Mitchell L, King M, Brillhart H, Goldstein A. 046 Cervical Ectropion may be a Cause of Desquamative Inflammatory Vaginitis. J Sex Med 2017;14:e366. 10.1016/j.jsxm.2017.04.048.

[24] Ma J, Kan Y, Zhang A, Lei Y, Yang B, Li P, et al. Female Sexual Dysfunction in Women with Non-Malignant Cervical Diseases: A Study from an Urban Chinese Sample. PLOS ONE 2015;10:e0141004. 10.1371/journal.pone.0141004.

[25] Rocha-Zavaleta L, Yescas G, Cruz RM, Cruz-Talonia F. Human papillomavirus infection and cervical ectopy. Int J Gynecol Obstet 2004;85:259–66. 10.1016/j.ijgo.2003.10.002.

[26] Sarkar PK, Steele PRM. Routine colposcopy prior to treatment of cervical ectopy: Is it worthwhile? J Obstet Gynaecol 1996;16:96–7. 10.3109/01443619609007753.

[27] Giroux V, Rustgi AK. Metaplasia: tissue injury adaptation and a precursor to the dysplasia–cancer sequence. Nat Rev Cancer 2017;17:594–604. 10.1038/nrc.2017.68.

[28] Rosenthal AN, Panoskaltsis T, Smith T, Soutter WP. The frequency of significant pathology in women attending a general gynaecological service for postcoital bleeding. BJOG Int J Obstet Gynaecol 2001;108:103–6. 10.1111/j.1471-0528.2001.00008.x.

[29] Lee A, Kim TH, Lee HH, Kim YS, Enkhbold T, Lee B, et al. Therapeutic Approaches to Atrophic Vaginitis in Postmenopausal Women: A Systematic Review with a Network Meta-analysis of Randomized Controlled Trials. J Menopausal Med 2018;24:1–10. 10.6118/jmm.2018.24.1.1.

[30] Kawakami PA, Velez-Montoya R, Castillejos-Chevez A, Remolina A, Garcia-Aguirre G, Rodriguez-Reyes A, et al. Safety And Efficacy Of Bismuth Subgallate As An Hemostatic Agent In An Animal Experimental Model. Invest Ophthalmol Vis Sci 2014;55:1510.

[31] Moscicki A-B, Ma Y, Holland C, Vermund SH. Cervical Ectopy in Adolescent Girls with and without Human Immunodeficiency Virus Infection. J Infect Dis 2001;183:865–70. 10.1086/319261.

[32] Jacobson DL, Peralta L, Graham NMH, Zenilman J. Histologic Development of Cervical Ectopy: Relationship to Reproductive Hormones. Sex Transm Dis 2000;27:252–8. 10.1097/00007435-200005000-00003.

[33] Wright K, Mohammed A, Salisu-Olatunji O, Kuyinu Y. Cervical Ectropion and Intra-Uterine Contraceptive Device (IUCD): a five-year retrospective study of family planning clients of a tertiary health institution in Lagos Nigeria. BMC Res Notes 2014;7:946.

