## Supplemental Table 1 for "Clinical Performance and Safety of Cerviron Vaginal Ovules in the Management of Cervical Lesions Postoperative Care: A National, Multicentric Study"

**Table 1.** The demography of the study participants.

| **Baseline characteristics** | **Experimental** | |
| --- | --- | --- |
|  | **n** | **%** |
| **Age (years): mean ± SD*** | 26 | 36.96 ± 10.28 |
| **Gender** |  |  |
| Female | 26 | 100.00 |
| **Ethnicity** |  |  |
| Caucasian | 26 | 100.00 |
| **Height (cm): mean ± SD** | 26 | 166.00 ± 5.51 |
| **Weight (kg): mean ± SD** | 26 | 65.31 ± 9.44 |
| **Subject in menopause** |  |  |
| Yes | 1 | 3.80 |
| No | 25 | 96.20 |
| **Tobacco consumption** |  |  |
| Yes | 6 | 23.10 |
| No | 20 | 76.90 |
| **Alcohol** |  |  |
| Yes | 2 | 7.69 |
| No | 23 | 88.46 |
| Unknown | 1 | 3.85 |
| **Regular Menstrual cycle** |  |  |
| Yes | 25 | 96.20 |
| No | 1 | 3.80 |

^*^ SD-Standard deviation.
