## Supplemental Table 2 for "Clinical Performance and Safety of Cerviron Vaginal Ovules in the Management of Cervical Lesions Postoperative Care: A National, Multicentric Study"

**Table 2.** The colposcopy examination and other clinical symptoms at baseline and after 90 days of treatment (final visit).

| **Colposcopy examination** | **Visits** | |
| --- | --- | --- |
|  | **Baseline** | **90 days** |
| **Normal** | 0 (00.00%) | 25 (100.00%) |
| **Abnormal** | 25 (100.00%) | 0 (00.00%) |
| **Total number of subjects** | 25 | 25 |
