## Supplemental Table 3 for "Clinical Performance and Safety of Cerviron Vaginal Ovules in the Management of Cervical Lesions Postoperative Care: A National, Multicentric Study"

**Table 3.** Analysis related to vaginal symptoms – comparison between visits.

| **Vaginal Symptoms** | **Baseline** | | **90 days** | | | **p-value** |
| --- | --- | --- | --- | --- | --- | --- |
|  | **Presence** | **Absence** | **Presence** | **Absence** |  | |
| **Primary Inflammation** | 24 (96.00%) | 1 (4.00%) | 0 (0.00%) | 25 (100.00%) | p < 0.001 | |
| **Secondary Inflammation** | 5 (20.00%) | 20 (80.00%) | 0 (0.00%) | 25 (100.00%) | p = 0.050* | |
| **Erosion** | 21 (84.00%) | 4 (16.00%) | 0 (0.00%) | 25 (100.00%) | p < 0.001 | |
| **Ulceration** | 17 (68.00%) | 8 (32.00%) | 0 (0.00%) | 25 (100.00%) | p < 0.001 | |
| **Colpitis** | 23 (92.00%) | 2 (8.00%) | 0 (0.00%) | 25 (100.00%) | p < 0.001 | |
| **Vaginal discharge** | 20 (80.00%) | 5 (20.00%) | 0 (0.00%) | 25 (100.00%) | p < 0.001 | |
| **Vaginal irritations** | 5 (20.00%) | 20 (80.00%) | 0 (0.00%) | 25 (100.00%) | p = 0.050* | |

p-value was obtained with the 2-sample test for equality of proportions with continuity correction *p-value was obtained with Fisher’s Exact Test
