## Supplemental Table 4 for "Clinical Performance and Safety of Cerviron Vaginal Ovules in the Management of Cervical Lesions Postoperative Care: A National, Multicentric Study"

**Table 4.** Degree of re-epithelialization between each visit.

| **Degree of re-epithelialization** | **Baseline** | **30 days** | **60 days** | **90 days** |
| --- | --- | --- | --- | --- |
| **Absent** | 18 (72.00%) | 0 (0.00%) | 0 (0.00%) | 0 (0.00%) |
| **Partial** | 7 (28.00%) | 24 (96.00%) | 3 (12.00%) | 0 (0.00%) |
| **Complete** | 0 (0.00%) | 1 (4.00%) | 22 (88.00%) | 25 (100.00%) |

^*^ SD-Standard deviation.
