## Supplemental Table 5 for "Clinical Performance and Safety of Cerviron Vaginal Ovules in the Management of Cervical Lesions Postoperative Care: A National, Multicentric Study"

**Table 5.** Means and Standard Deviations of each symptom between visits.

| **Measure** | **Baseline** | **30 days** | **60 days** | **90 days** | **p-value** |
| --- | --- | --- | --- | --- | --- |
| **Bleeding** | 1.44 ± 1.16 | 0.56 ± 0.71 | 0.04 ± 0.20 | 0.00 ± 0.00 | p < 0.001 |
| **Malodor** | 1.52 ± 1.33 | 0.56 ± 0.71 | 0.00 ± 0.00 | 0.00 ± 0.00 | p < 0.001 |
| **Dysuria** | 1.40 ± 1.35 | 0.80 ± 1.19 | 0.16 ± 0.62 | 0.20 ± 1.00 | p < 0.001 |
| **Dyspareunia** | 1.68 ± 1.57 | 0.68 ± 0.85 | 0.00 ± 0.64 | 0.00 ± 0.00 | p < 0.001 |
| **Pain** | 1.60 ± 1.32 | 0.56 ± 0.65 | 0.00 ± 0.00 | 0.00 ± 0.00 | p < 0.001 |
| **Leucorrhea** | 2.08 ± 1.38 | 0.56 ± 0.65 | 0.00 ± 0.00 | 0.00 ± 0.00 | p < 0.001 |

*Note*. Mean and SD are presented by the form μ and the SD

p value is obtained using the Kruskall-Wallis nonparametric test.
